# ClinStigma: A Clinician-Guided Framework for Developing Synthetic Benchmarks of Contextual Stigmatizing Language in Clinical Documentation

**DOI:** 10.64898/2026.10.10.26365233

**Authors:** Joseph A. Izzo, Adrienne M. McIntyre, Jacklyn Nguyen, Dennis Bashaw, Caprice A. Torrance, Jennifer Foster

## Abstract

**Objective:** Large language models (LLMs) are increasingly used to generate, summarize, and evaluate clinical documentation, yet standardized methods for evaluating nuanced, context-dependent clinical language remain limited. Existing approaches rely primarily on annotated clinical corpora or keyword-based methods, restricting reproducibility, public dissemination, and systematic comparison across models. We present ClinStigma, a clinician-guided framework for constructing, annotating, and evaluating synthetic benchmarks of contextual stigmatizing language, demonstrated through ClinStigma-SUD, its inaugural implementation for substance use disorder documentation.

**Materials and Methods:** Candidate clinical sentences were generated using a frontier language model and iteratively reviewed according to a clinician-developed annotation rubric refined through expert review in stigmatizing language. The resulting benchmark intentionally incorporates explicit stigmatizing, neutral, and contextually ambiguous (“gray-zone”) documentation requiring clinical reasoning rather than keyword recognition. Three locally deployed open-weight LLMs were evaluated using zero-shot inference, followed by external evaluation using independently annotated clinical documentation from MIMIC-IV.

**Results:** ClinStigma-SUD consists of 500 clinician-adjudicated synthetic clinical documentation sentences. Inter-rater agreement was 98.8% across the full benchmark (Cohen’s κ = 0.976) and 94.0% within the intentionally ambiguous subset (κ = 0.880). Under zero-shot prompting, binary accuracy ranged from 84.8% to 91.8% across evaluated models, while F1 for the ambiguous (gray) class ranged from 0.038 to 0.423. Zero-shot external evaluation on 3,716 annotated examples from the independently annotated Harrigian MIMIC-IV corpus yielded 84.4% accuracy (84.4% balanced accuracy; macro F1 0.824), against a 69.9% majority-class baseline. Stigmatizing-class F1 was 0.879 in the substance-related subgroup and 0.883 in the full external corpus.

**Discussion and Conclusion:** ClinStigma establishes a reproducible clinician-guided framework for developing synthetic evaluation benchmarks for contextual clinical language. By combining structured clinical expertise, explicit annotation guidance, synthetic benchmark generation, and external evaluation, the framework enables open, privacy-preserving evaluation of language models while providing a scalable foundation for benchmark development across additional clinical specialties, including obstetrics, emergency medicine, and mental health.

## 1. Introduction

Stigmatizing language in electronic health records (EHRs) contributes to bias and may adversely influence patient experience, clinical decision-making, and engagement in care for individuals with substance use disorder (SUD).^1–3^ Language that subtly communicates disbelief, moral judgment, or diminished credibility has been shown to influence subsequent clinicians who review the medical record, propagating bias throughout the continuum of care.^1^

Traditional approaches to identifying stigmatizing language have relied on manual chart review, keyword lexicons, or annotation of real-world clinical corpora.^2,3^ Although these methods have advanced understanding of stigma in clinical documentation, they present practical challenges for reproducibility, scalability, and public dissemination. Access to authentic clinical documentation is frequently restricted by privacy regulations, while keyword-based approaches often fail to capture contextual distinctions involving attribution, clinical hedging, behavioral framing, and other nuanced linguistic phenomena.

Recent advances in large language models (LLMs) have demonstrated promising performance for automated detection of stigmatizing language in clinical documentation, although performance varies substantially with model configuration, prompting strategy, and clinical note context.^4–7^ However, evaluation remains constrained by the absence of openly distributable, clinician-guided benchmarks capable of systematically assessing contextual clinical reasoning. Existing benchmark development has largely focused on annotation of real clinical corpora rather than reproducible methodologies for constructing and assessing synthetic evaluation resources.^8^

To address this gap, we present ClinStigma, a clinician-guided framework for developing synthetic benchmarks for contextual stigmatizing language in clinical documentation. ClinStigma integrates literature-informed taxonomy development, structured annotation rubric design, clinician adjudication, synthetic benchmark generation, and external evaluation into a reproducible methodology for benchmark construction. We demonstrate the framework through **ClinStigma-SUD**, the inaugural implementation of the framework, and evaluate three locally deployed open-weight language models before externally evaluating benchmark findings using previously annotated clinical documentation from MIMIC-IV.^8,9^

## 2. Materials and Methods

### 2.1 Overview

We developed **ClinStigma**, a clinician-guided framework for constructing synthetic evaluation benchmarks for contextual stigmatizing language in clinical documentation. The framework was designed to transform clinician expertise into a reproducible methodology for creating openly distributable benchmarks that enable systematic evaluation of large language models (LLMs) without requiring access to protected health information (PHI) or restricted clinical corpora.

Framework development and evaluation consisted of five core components: (1) literature-informed taxonomy and annotation rubric development, (2) synthetic benchmark generation, (3) clinician review and adjudication, (4) local LLM evaluation, and (5) external evaluation using annotated real-world clinical documentation (Figure 1).

**Figure 1.**
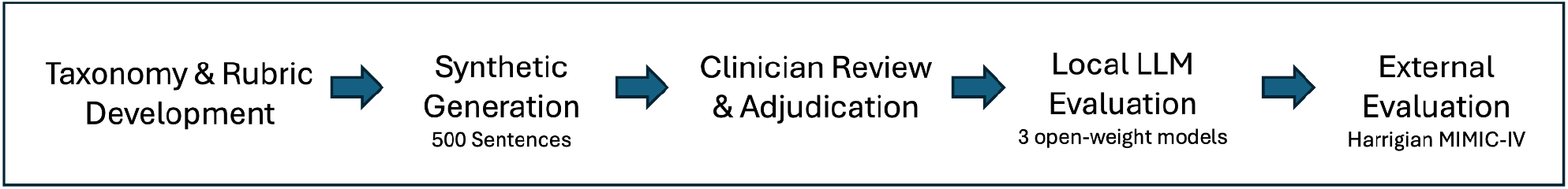
ClinStigma framework for clinician-guided synthetic benchmark development and evaluation.

Substance use disorder (SUD) was selected as the inaugural implementation because it represents a clinically important domain in which distinctions between objective documentation and stigmatizing language frequently depend on contextual clinical judgment rather than isolated lexical features.

### 2.2 Literature-Informed Taxonomy Development

An initial taxonomy of stigmatizing clinical language was developed through clinician domain expertise, review of prior literature on stigmatizing clinical documentation and recommended substance use disorder terminology, and iterative review of candidate benchmark examples.^3,10–14^ Categories included identity-based labeling, credibility framing, behavioral framing, clinician attribution, documentation hedging, refusal language, moralized terminology, and context-dependent documentation. Particular emphasis was placed on language requiring contextual interpretation rather than recognition of predefined keywords. The taxonomy and associated benchmark categories were iteratively refined during benchmark design and early candidate generation before construction of the final benchmark.

### 2.3 Annotation Rubric Development

A structured clinician annotation rubric was developed to operationalize contextual stigmatizing language for benchmark construction. Rather than defining stigma using isolated words or phrases, the rubric established explicit decision rules for classifying documentation as stigmatizing or neutral and identified circumstances in which classification was context dependent or potentially ambiguous.

The rubric included operational definitions for multiple linguistic domains, including:

- identity-first versus person-first language
- credibility framing
- clinician attribution
- behavioral framing
- refusal language
- documentation hedging
- judgmental phrasing
- moralized terminology

Special attention was given to “gray-zone” documentation, defined as clinical language in which interpretation depends on surrounding context, provider intent, attribution, or clinical reasoning rather than vocabulary alone.

The rubric was developed iteratively by a board-certified Addiction Medicine physician and refined through expert review by a Doctor of Nursing Practice (DNP)-prepared clinician with expertise in stigmatizing language and clinical communication. Selected examples of decision rules from the annotation rubric are shown in Table 1.

**Table 1.** Selected examples of decision rules from the ClinStigma annotation rubric used during clinician adjudication.

| Term | Stigmatizing When |
| --- | --- |
| Refused | Frames reasonable behavior as obstinance |
| Claims | Implies clinician disbelief |
| Attribution | Provider-sourced; not lay family |
| Despite | Judgmental framing rather than chronology |
| Clean/Dirty | Frames test results using moralized language |

### 2.4 Synthetic Benchmark Construction

Synthetic benchmark construction was designed to generate clinically realistic documentation while avoiding inclusion of protected health information.

Candidate sentences were generated using Claude Sonnet 4.6.^15^ Rather than relying on a single generation prompt, benchmark construction consisted of repeated interactions in which the model generated candidate clinical documentation for prespecified generation subcategories.

Generated examples were reviewed by the study investigators and revised, discarded, or regenerated as needed to improve clinical realism and consistency with the intended annotation criteria. The generation emphasized contextual diversity rather than lexical diversity.

Following iterative benchmark design, the final generation structure comprised 12 prespecified subcategories reflecting explicit stigmatizing language, neutral contrast documentation, and contextually ambiguous scenarios. These included four stigmatizing subcategories (identity labels, behavioral framing, distrust language, and moral judgment), three neutral contrast subcategories (person-first language, clinical diagnostic terminology, and factual behavioral documentation), and five gray-zone subcategories (context-dependent refusal, quoted legacy diagnostic terminology, clinical hedging, third-party attributed language, and behavioral documentation without judgment).

An initial benchmark of 500 candidate sentences was produced consisting of:

- 200 explicitly stigmatizing examples
- 200 clearly neutral examples
- 100 intentionally ambiguous (“gray-zone”) examples

Gray-zone examples were specifically designed to evaluate contextual reasoning beyond keyword recognition. For example, language such as *“patient claims*…*”* may represent either objective attribution or implied clinician disbelief depending upon surrounding documentation context.

### 2.5 Clinician Adjudication

All benchmark candidates underwent clinician review by a board-certified Addiction Medicine physician using the structured annotation rubric.

Each sentence was evaluated according to overall contextual meaning rather than isolated lexical features. Candidate examples were accepted, revised, relabeled, or excluded based on consistency with rubric definitions. Gray-zone examples received particular attention because they represented the primary objective of benchmark development: evaluating contextual clinical reasoning rather than explicit terminology recognition.

Following adjudication, the finalized benchmark consisted of 251 stigmatizing and 249 neutral examples. Gray-zone scenarios remained embedded within both outcome classes according to final clinician judgment rather than existing as an independent output category.

A second Doctor of Nursing Practice (DNP)-prepared clinician with expertise in stigmatizing language and clinical communication independently reviewed the finalized benchmark using the same annotation framework. Inter-rater reliability was assessed across the complete 500-item benchmark and separately within the 100 intentionally ambiguous examples. The independent reviewer also documented the clinical rationale for disagreements to characterize sources of residual contextual ambiguity.

### 2.6 Benchmark Characteristics

The finalized benchmark, ClinStigma-SUD v1.0, contains 500 clinician-adjudicated synthetic clinical sentences representing a broad spectrum of contextual stigmatizing language encountered in substance use disorder documentation.

Unlike lexicon-based resources, the benchmark intentionally incorporates attribution, credibility language, behavioral framing, documentation hedging, refusal language, and context-dependent clinical reasoning. The benchmark was designed as an openly distributable evaluation resource accompanied by the ClinStigma taxonomy, annotation rubric, dataset documentation, and version-controlled releases to facilitate extension into additional clinical specialties.

### 2.7 Model Evaluation

To characterize benchmark behavior, three contemporary open-weight LLMs were evaluated using zero-shot prompting without fine-tuning, retrieval augmentation, or parameter adaptation:

- Gemma 4 31B (bf16)^16^
- Llama 3.3 70B (8-bit)^17^
- Qwen3.6-35B A3B^18^

Inference was performed locally on Apple Silicon hardware using the MLX inference framework.^19^ All evaluations used zero-shot prompting with temperature set to 0 and without fine-tuning, retrieval augmentation, or parameter adaptation. Models were evaluated under separate binary and ternary zero-shot classification tasks. For binary classification, models classified each sentence as stigmatizing or neutral using the final clinician-adjudicated reference labels. For ternary classification, models classified sentences as stigmatizing, neutral, or gray, with the 100 intentionally ambiguous examples serving as the gray-zone reference subset.

Performance was assessed using overall accuracy, precision, recall, F1-score, confusion matrices, and benchmark subcategory-specific accuracy.

### 2.8 External Evaluation on Annotated Clinical Documentation

External evaluation was performed using the previously annotated Harrigian MIMIC-IV corpus.^8,9^ The source corpus contained 5,043 annotations. After removing 1,137 annotations labeled as difficult or excluded, 3,906 annotations across 3,664 clinical notes remained for context reconstruction.

Source annotations provided annotated keywords and character offsets rather than complete sentences. For each annotation, a symmetric 10-word context window was reconstructed around the annotated keyword. Because reported character offsets did not always align with the local MIMIC-IV note text, the annotated keyword was searched within a ±500-character window around the reported offset. An additional 190 annotations for which the keyword could not be recovered within this window were excluded, yielding a final external evaluation set of 3,716 examples.

Source labels were mapped to binary reference categories, with negative and disbelief annotations classified as stigmatizing and positive and neutral annotations classified as neutral. Source keyword categories were summarized as credibility and distrust (adamant), compliance, and descriptors (other). All adamant annotations carried disbelief labels.

Gemma 4 31B was evaluated using zero-shot classification with an external-evaluation prompt adapted to the Harrigian annotation structure. The model was provided with the reconstructed clinical context, annotated keyword, and keyword category. The prompt also included guidance for linguistic categories represented in the Harrigian corpus but not explicitly covered by the ClinStigma-SUD benchmark prompt, including appearance and behavioral descriptors, credibility or exaggeration framing, and concessive constructions such as “despite.” Overall accuracy, balanced accuracy, macro F1, and class-specific precision, recall, and F1-score were calculated.

A substance-related subgroup was defined using hospitalizations with any listed ICD-10-CM diagnosis from F10 through F19, excluding F17, yielding 337 evaluable examples. Performance within this subgroup was compared with performance across the full external evaluation corpus.

### 2.9 Statistical Analysis

Overall classification accuracy, precision, recall, and F1-score were calculated for each evaluated model. Performance was additionally summarized across benchmark generation subcategories to identify strengths and limitations in contextual language understanding.

Because contextual ambiguity represented the primary motivation for benchmark development, analyses emphasized model behavior within clinician-adjudicated gray-zone scenarios, where distinctions between objective documentation and stigmatizing language depend on contextual clinical reasoning rather than lexical detection.

Inter-rater reliability was assessed using observed percent agreement and Cohen’s kappa (κ), calculated for the complete benchmark and separately for the intentionally ambiguous subset. Disagreements identified during independent review were examined alongside reviewer-provided rationales to characterize sources of annotation disagreement.

## 3. Results

### 3.1 Inter-Rater Reliability

Independent clinician review demonstrated high agreement across the full ClinStigma-SUD benchmark, with 98.8% observed agreement and Cohen’s κ = 0.976. Agreement remained high within the 100 intentionally ambiguous gray-zone examples (94.0%; κ = 0.880). Reviewer-documented disagreements reflected differences in contextual interpretation within deliberately ambiguous scenarios rather than disagreement regarding explicit stigmatizing terminology.

### 3.2 Zero-Shot Model Performance

Three contemporary open-weight large language models were evaluated on the ClinStigma-SUD benchmark using zero-shot prompting. Gemma 4 31B achieved the highest overall binary classification accuracy (91.8%), followed by Llama 3.3 70B (88.6%) and Qwen3.6-35B A3B (84.8%) (Figure 2). Although overall performance was high across all evaluated models, differences emerged in the handling of contextually ambiguous clinical language.

**Figure 2.**
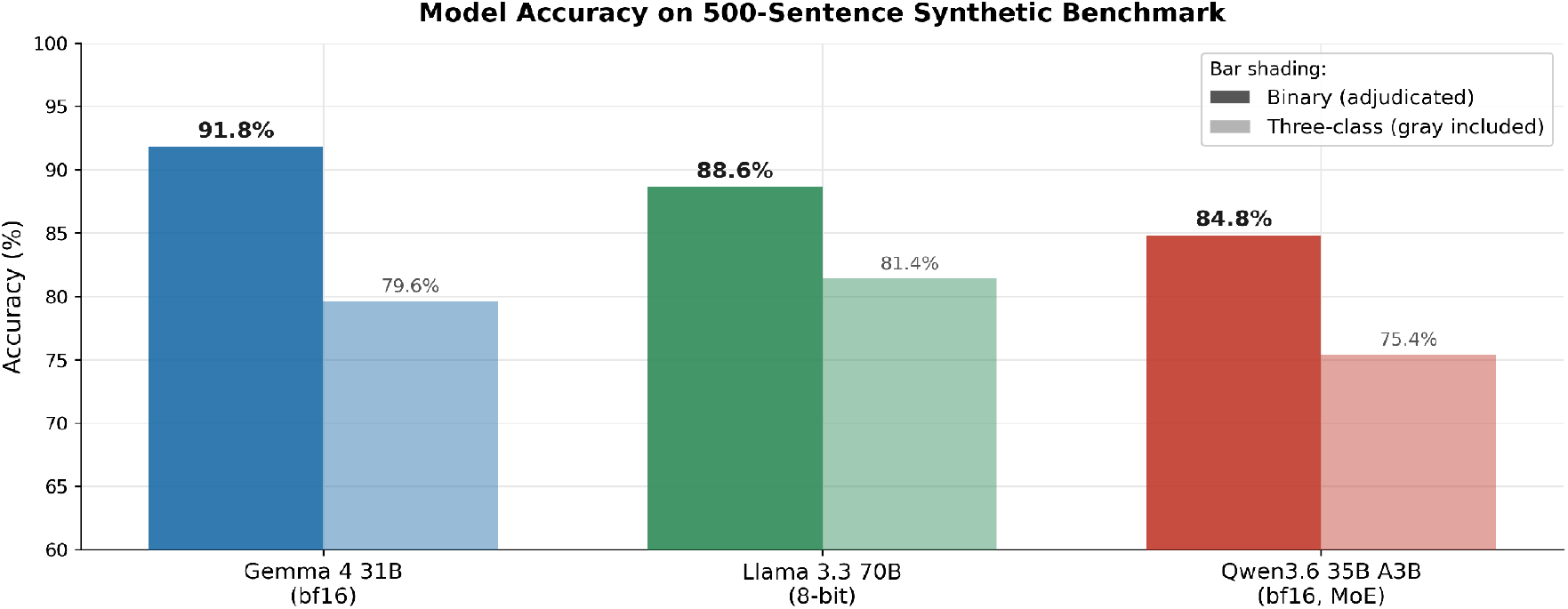
Zero-shot model performance on the ClinStigma-SUD benchmark. Binary accuracy reflects classification against final clinician-adjudicated stigmatizing versus neutral reference labels; three-class accuracy reflects classification as stigmatizing, neutral, or gray.

### 3.3 Performance Across Benchmark Subcategories

Performance varied substantially across benchmark subcategories (Figure 3). Models demonstrated high or near-ceiling performance across several categories characterized by relatively explicit lexical cues, including person-first language, identity labels, and clinical diagnostic terminology. Greater variability emerged in categories requiring contextual interpretation, including refusal language, clinician attribution, behavioral framing, and quoted legacy terminology. The largest performance differences occurred within context-dependent refusal language, where binary accuracy ranged from 12% to 40% across evaluated models. Overall, errors clustered within categories requiring contextual clinical reasoning rather than explicit lexical recognition.

**Figure 3.**
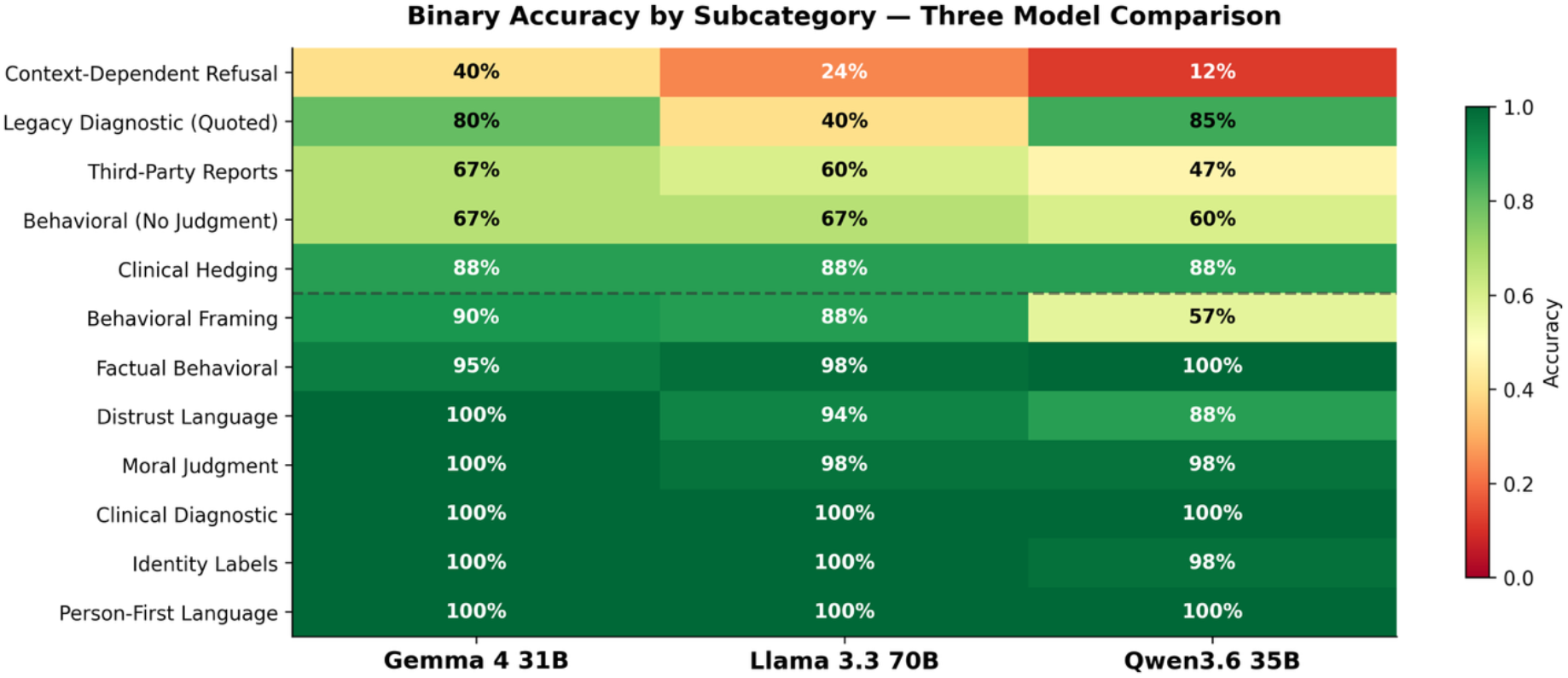
Binary classification accuracy across ClinStigma-SUD benchmark generation subcategories. The upper five rows represent intentionally ambiguous gray-zone subcategories.

### 3.4 Model Behavior Under Contextual Ambiguity

Although binary classification performance remained high, substantial differences emerged when evaluating clinician-adjudicated gray-zone examples (Figure 4). Llama 3.3 70B demonstrated the highest gray-class F1 (0.423) whereas Gemma 4 31B and Qwen3.6-35B A3B more frequently assigned definitive classifications (gray F1 = 0.215 and 0.038, respectively).

**Figure 4.**
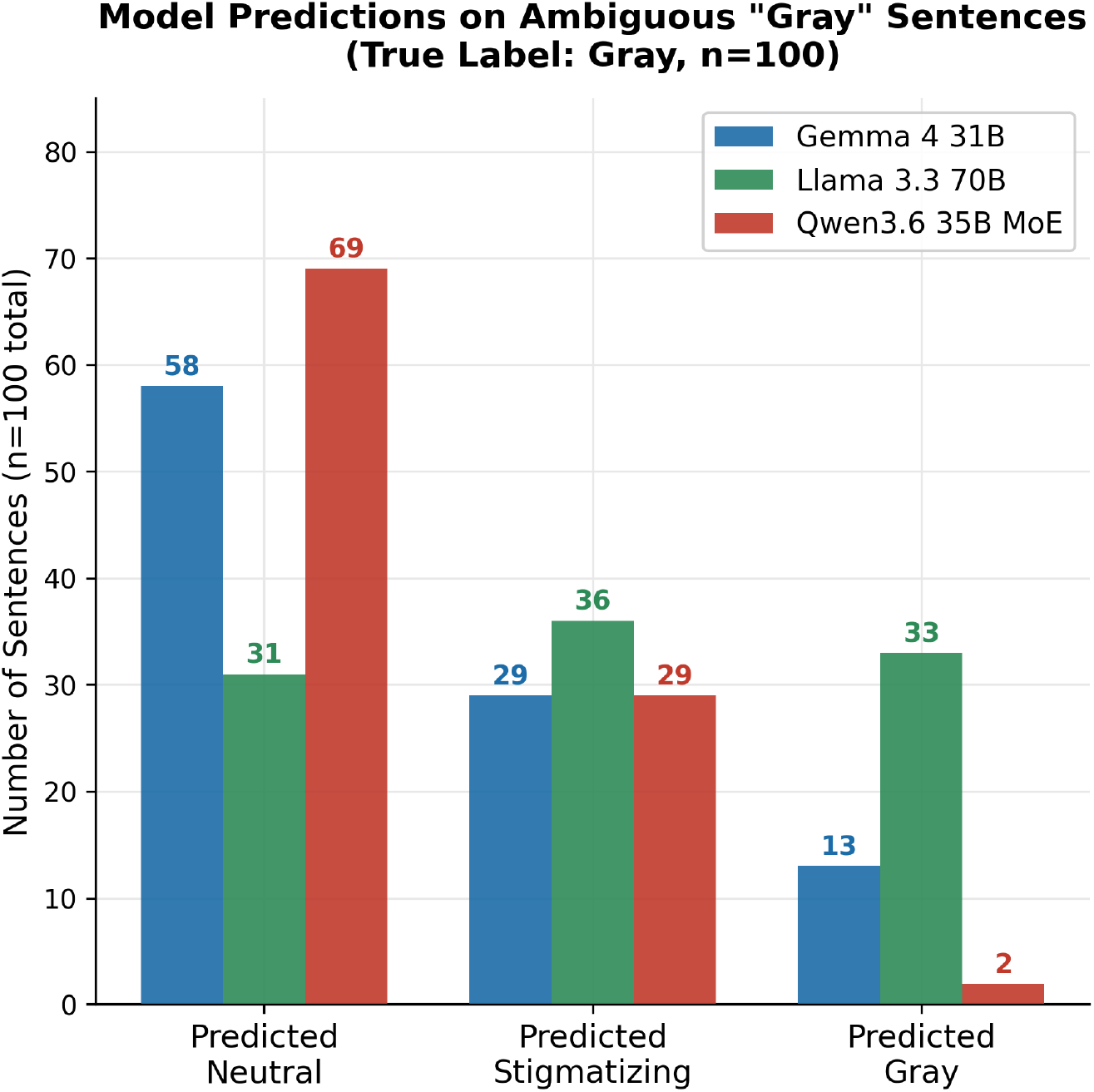
Model predictions on intentionally ambiguous gray-zone examples.

Inspection of model predictions demonstrated distinct classification behavior under ambiguity. Gemma preferentially assigned neutral classifications in ambiguous scenarios, whereas Qwen rarely assigned gray classifications despite similar binary accuracy. Llama more frequently selected the gray category, indicating greater sensitivity to contextual ambiguity under this task. Collectively, these findings indicate that contextual framing, rather than explicit stigmatizing terminology, represents the principal challenge for automated stigma detection.

### 3.5 External Evaluation

The full external evaluation included 3,716 annotated clinical examples, of which 2,596 were stigmatizing and 1,120 neutral. Gemma 4 31B achieved 84.4% overall accuracy and 84.4% balanced accuracy, compared with a 69.9% majority-class accuracy baseline. Macro F1 was 0.824. For stigmatizing examples, precision was 0.926, recall was 0.844, and F1 was 0.883. For neutral examples, precision was 0.700, recall was 0.845, and F1 was 0.766. Errors occurred in both directions, with 405 stigmatizing examples classified as neutral and 174 neutral examples classified as stigmatizing (Figure 5).

**Figure 5.**
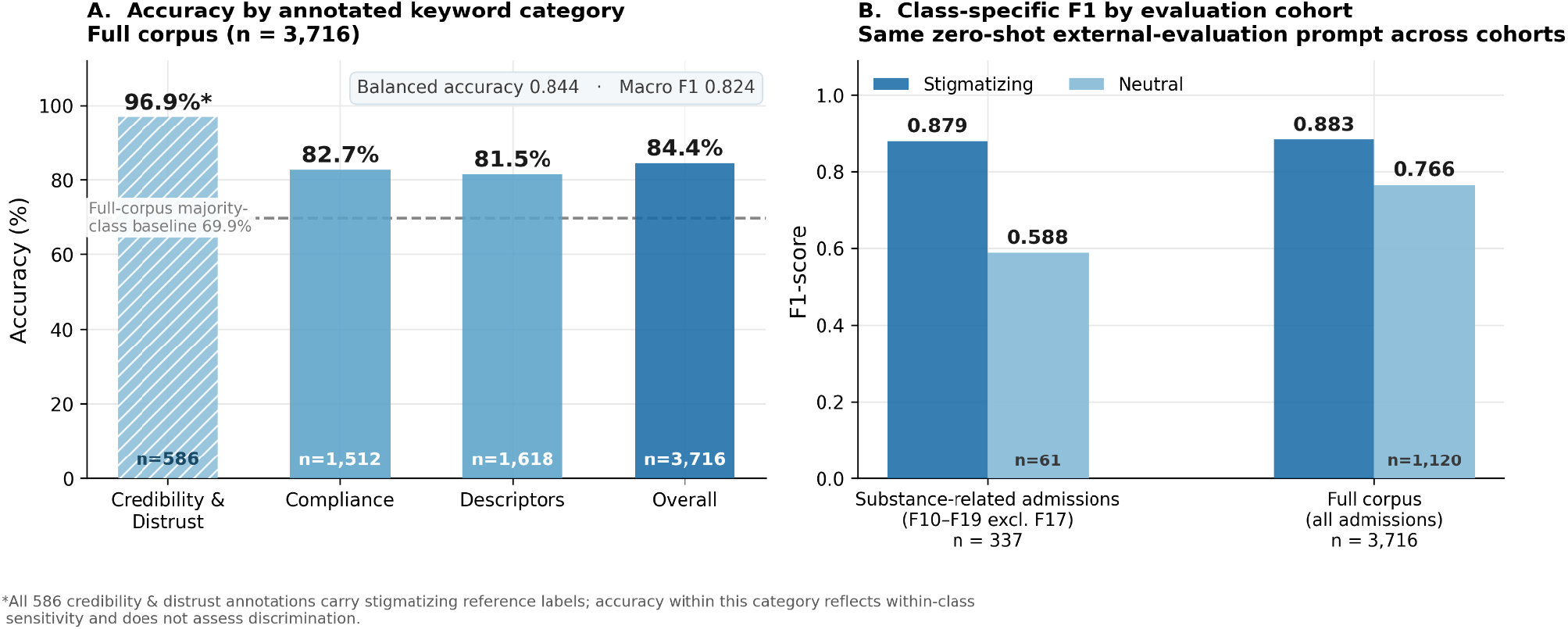
External evaluation of Gemma 4 31B on annotated Harrigian MIMIC-IV clinical documentation.

Performance was similar across source categories containing both reference classes. Accuracy was 82.7% for compliance language and 81.5% for descriptor language. Accuracy was 96.9% for credibility and distrust language; however, all 586 examples in this category were disbelief annotations and therefore carried stigmatizing reference labels. This category therefore reflects within-class sensitivity rather than discrimination between stigmatizing and neutral documentation.

Within the substance-related subgroup including alcohol and other substance-related disorders (n = 337), overall accuracy was 81.3%, compared with an 81.9% majority-class accuracy baseline. Balanced accuracy was 78.4% and macro F1 was 0.734. Stigmatizing-class F1 was similar in the substance-related subgroup and full external corpus (0.879 and 0.883, respectively). Neutral-class F1 was 0.588 in the substance-related subgroup and 0.766 in the full corpus; these estimates were based on 61 and 1,120 neutral reference examples, respectively.

## 4. Discussion

ClinStigma demonstrates that clinician-guided synthetic benchmarks can provide a reproducible, privacy-preserving approach for evaluating contextual stigmatizing language in clinical documentation. Rather than relying exclusively on restricted clinical corpora or keyword-based methods, the framework integrates literature-informed taxonomy development, structured annotation guidance, clinician adjudication, and external evaluation to produce openly distributable evaluation resources.

Although all evaluated models demonstrated high performance for explicit stigmatizing language, substantial differences emerged when interpreting contextually ambiguous documentation. Errors clustered within clinician-adjudicated gray-zone examples, suggesting that contextual clinical reasoning, not lexical recognition, remains the principal challenge for automated stigma detection. These findings emphasize the importance of evaluating model behavior under uncertainty in addition to traditional accuracy metrics.

High inter-rater agreement provides additional evidence that this difficulty does not simply reflect instability of the reference labels. Clinicians applying the structured annotation framework demonstrated near-complete agreement across the benchmark and high agreement even within deliberately ambiguous examples, whereas evaluated models demonstrated substantially greater difficulty classifying deliberately ambiguous examples. This distinction supports the use of clinician-guided gray-zone examples as a targeted evaluation of contextual language understanding.

External evaluation provided complementary evidence that the distinctions represented in ClinStigma-SUD are detectable in authentic clinical documentation. Across 3,716 independently annotated clinical examples, Gemma 4 31B achieved 84.4% overall accuracy and 84.4% balanced accuracy under zero-shot classification, compared with a 69.9% majority-class accuracy baseline. Stigmatizing-class F1 was similar in the substance-related subgroup and the full external corpus (0.879 and 0.883, respectively), suggesting that detection of stigmatizing language was not specific to substance-related documentation.

ClinStigma is intended as a reproducible methodology rather than a single benchmark. The framework can be instantiated within a clinical domain by defining a domain-specific taxonomy and annotation rubric, generating and adjudicating synthetic examples, benchmarking model behavior, and, where available, evaluating performance against independently annotated clinical text.

## 5. Limitations

Several limitations should be considered. First, synthetic benchmark examples, despite clinician review and external evaluation, cannot fully reproduce the complexity and longitudinal context of authentic clinical documentation. Benchmark generation relied on a single language model, which may introduce model-specific stylistic patterns not fully representative of naturally occurring clinical documentation. Second, external evaluation was limited to a single annotated clinical corpus and one externally evaluated language model. Context-window reconstruction required approximation where source annotation offsets did not align with local MIMIC-IV note text. The external corpus also remained class imbalanced, and the credibility and distrust category contained only stigmatizing reference labels. Third, although independent clinician review demonstrated high inter-rater agreement, the annotation framework was evaluated by a limited number of clinical experts and may not capture the full range of perspectives regarding context-dependent stigmatizing language. Finally, models were evaluated using zero-shot prompting without fine-tuning or retrieval augmentation, and benchmark performance may differ following domain-specific adaptation.

## 6. Conclusion

We present ClinStigma, a clinician-guided framework for constructing synthetic evaluation benchmarks of contextual stigmatizing language in clinical documentation, together with ClinStigma-SUD, its inaugural implementation. By combining structured clinical expertise, synthetic benchmark construction, clinician adjudication, and external evaluation, ClinStigma provides an openly distributable, privacy-preserving methodology for evaluating large language models on nuanced clinical language tasks. This framework establishes a foundation for future benchmark development across additional clinical domains while supporting reproducible evaluation of emerging language models in a controlled, privacy-preserving setting prior to application to patient data.

## Data Availability

The ClinStigma-SUD v1.0 benchmark, annotation rubric, evaluation prompts, and accompanying documentation will be made publicly available at https://github.com/izzomd/ClinStigma and https://huggingface.co/datasets/ClinStigma/ClinStigma-SUD following release of this preprint. The benchmark consists entirely of synthetic clinical documentation and contains no protected health information. External evaluation used previously released annotations from the Harrigian MIMIC-IV corpus, available to credentialed users through PhysioNet under its data use agreement. Those annotations, along with the context windows and model outputs derived from restricted MIMIC-IV data, may not be publicly redistributed by the authors.

https://github.com/izzomd/ClinStigma

## Ethics and Data Use

This study received an exemption determination from the institutional review board. ClinStigma-SUD consists entirely of synthetic clinical documentation and does not contain patient data or protected health information. External evaluation was performed using deidentified MIMIC-IV data and previously released annotations obtained through PhysioNet under the applicable credentialing and data-use requirements.

